# Allostatic load modifies neuropsychiatric risk following traumatic brain injury

**DOI:** 10.64898/2026.06.21.26356173

**Authors:** Tadeusz H. Wroblewski, Peter B. Barr, Tim B. Bigdeli, Ernest J. Barthélemy

## Abstract

**Importance:** Outcomes following traumatic brain injury (TBI) vary substantially, with a subset of individuals experiencing neuropsychiatric morbidity and worse prognosis. Exposure to psychosocial and environmental stressors may be an important, yet understudied, modifier of TBI trajectory. Allostatic load (AL) represents the cumulative physiological burden of chronic stress and provides a useful framework for evaluating pre-injury vulnerability.

**Objective:** To assess the relationship between pre-injury AL burden and risk of mortality and incident neuropsychiatric diagnosis following TBI.

**Design, Setting, and Participants:** This cohort study leveraged electronic health record, survey, and laboratory data from the All of Us Research Program, version 8. Participants aged 18 years or older enrolled between May 6, 2018, and October 1, 2023, were queried for TBI diagnosis using clinical diagnostic codes. Data were analyzed between November 11, 2024, and January 7, 2026.

**Exposure:** The physiological burden of pre-injury chronic stress exposure was estimated using an AL index (pALI) derived from anthropometric and laboratory biomarkers collected before index TBI.

**Main Outcomes and Measures:** Post-TBI mortality and incident neuropsychiatric diagnosis clusters. Mortality risk was assessed using Cox proportional hazards models (hazard ratio [HR] with 95% CI), and risk of incident neuropsychiatric diagnosis was modeled using competing-risk regression with death as a competing event (sub-distribution HR with 95% CI).

**Results:** The primary cohort included 4,552 individuals with an established TBI diagnosis and sufficient biomarker data to estimate pALI. The pALI measure differed across sociodemographic groups and was positively correlated with perceived stress (*r*=.08, *p*=.002). Higher pALI was associated with increased post-TBI mortality risk (adjusted HR=1.71; 95%CI, 1.36-2.14).

Elevated pALI was also associated with greater risk of incident post-traumatic stress disorder (PTSD; adjusted HR=1.28; 95%CI, 1.10-1.50) and sleep disorder (adjusted HR=1.42 95%CI, 1.29-1.57) diagnoses.

**Conclusions and Relevance:** Higher pre-injury ALI was associated with increased risk of mortality and select neuropsychiatric outcomes following TBI, suggesting that AL burden may shape post-injury trajectories. Pre-injury chronic stress exposure and underlying stress biology may represent underrecognized determinants of vulnerability and resilience in brain injury recovery.

**KEY POINTS:** *Question:* Is the physiological burden of chronic stress, estimated by allostatic load (AL), associated with worse outcomes following traumatic brain injury (TBI)?

*Findings:* Among 4,552 All of Us participants with TBI, higher pre-injury AL was associated with increased mortality risk and greater incidence of post-traumatic stress disorder and sleep disorder diagnosis following injury.

*Meaning:* These findings suggest that the cumulative physiological burden of chronic stress may be an underrecognized modifier of TBI recovery and outcomes that could inform brain trauma risk stratification and prognostication.

## INTRODUCTION

Trajectories following traumatic brain injury (TBI) are marked by substantial heterogeneity, with a subset of individuals experiencing persistent cognitive impairment, reduced quality of life, and increased risk for neuropsychiatric disease.^1–3^ Biopsychosocial and environmental (BPSE) modifiers^4,5^ contribute to this variability and have recently been formalized within the <u>C</u>linical-<u>B</u>iomarker-Imaging-<u>M</u>odifier (CBI-M) framework established by the National Institute of Neurological Disorders and Stroke (NINDS) TBI Classification and Nomenclature Initiative.^6^ BPSE factors – including access to medical care, medical and mental health burden, and other structural determinants of health – contribute to chronic psychosocial stress ^4,7–9^. Thus, chronic exposure to psychosocial and environmental stressors may represent a shared mechanism through which the Modifier pillar contributes to variability in TBI outcomes.^3,10^

Chronic stress engages stress-responsive systems which have downstream effects driving neurological morbidity and adverse health outcomes.^11,12^ Prior evidence suggest a role of pre-existing chronic stress in mediating depressive, somatic, and fatigue-related symptoms following TBI.^13–15^ TBI itself constitutes a profound psychological and physiological stressor,^3^ with effects extending beyond the initial insult through dynamic processes shaped by environmental and genetic influences.^16–18^ The cumulative biological consequence of sustained stress has been conceptualized as allostatic load (AL),^19^ a framework which describes the physiological “wear and tear” resulting from repeated and dysregulated adaptive responses.^20^ Furthermore, biological processes implicated in allostatic overload overlap with secondary mechanisms of TBI, including persistent neuroinflammation, oxidative stress, and cerebrovascular dysfunction.^10,21^ In this context, AL can provide a unifying framework, linking environmental psychosocial stress exposure to underlying stress biology and post-TBI trajectories.

The combined effects of chronic stress exposure and brain trauma warrant further investigation into how these insults may compound to impact post-TBI trajectory. We have previously operationalized TBI and incident neuropsychiatric sequalae phenotypes in the longitudinal All of Us (AoU) Research Program cohort.^22,23^ In the present study, we leverage this framework to examine the association of pre-injury cumulative stress burden with post-TBI neuropsychiatric morbidity and mortality using a time-sensitive AL index constructed from laboratory and anthropometric measures.

## METHODS

### Conceptual design

This cohort study evaluated how the physiological burden of pre-injury chronic stress impacts prognosis following brain trauma [Figure 1]. To approximate a time-dependent measure of physiological stress exposure, an AL index was operationalized using established biomarkers^24–26^ with measurements restricted to those obtained prior to the index injury. We then assessed associations between pre-injury AL index (pALI) and post-TBI outcomes, including mortality and risk of incident neuropsychiatric diagnoses. Incident diagnoses were defined as the first recorded occurrence of a given neuropsychiatric diagnosis across available EHR. This study was conducted in accordance with the AoU Code of Conduct and the State University of New York Downstate Health Sciences University Institutional Review Board.

**Figure 1.**
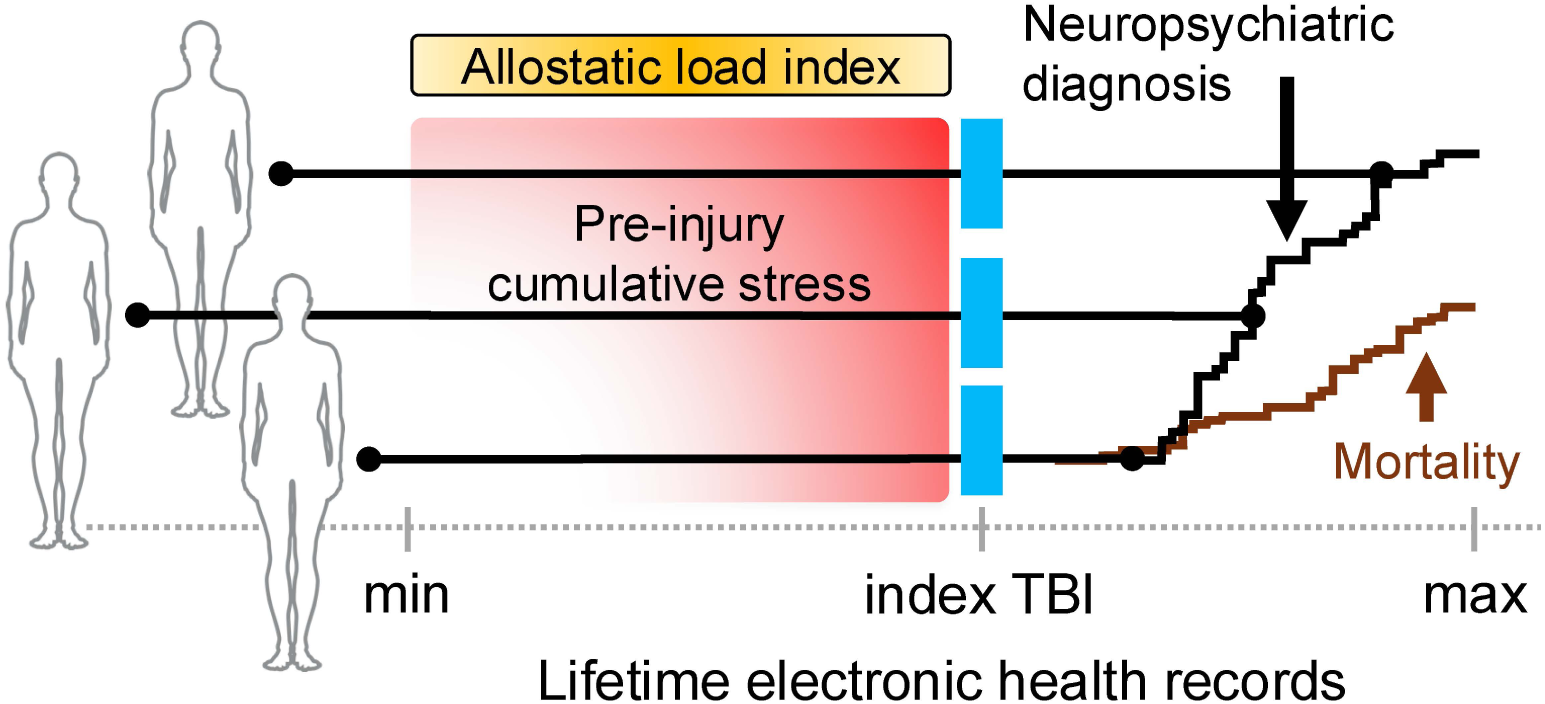
Conceptual diagram depicting the assessment of pre-injury allostatic load and traumatic brain injury outcomes. Cumulative stress prior to index TBI was estimated using a pre-injury allostatic load index (pALI) derived from blood-based and physical biomarkers. The pre-injury average of each marker was used to estimate pALI. The impact of pALI was then assessed on incident phecode-derived neuropsychiatric diagnoses and mortality.

### Study population

The AoU Research Program integrates longitudinal electronic health record (EHR) data, survey responses, and physical and laboratory measurements from participants across the United States,^27^ with an emphasis on increasing representation of populations historically underrepresented in biomedical research. Participants enroll, provide informed consent, complete surveys, and link EHR data through an online portal (joinallofus.org). Participants 18 years or older at the time of enrollment, between May 6, 2018, and October 1, 2023 (v8; *N*=393,601), were queried for TBI diagnosis codes.

We applied our previously described TBI definition,^22^ derived from *International Classification of Diseases, Ninth Revision* (ICD-9) and *Tenth Revision Clinical Modification* (ICD-10CM) diagnosis codes from the Centers for Disease Control and Prevention,^28^ Department of Defense,^29^ and clinical review from the consulting neurosurgeon (E.J.B.) [Table S1]. Participants required at least one documented TBI diagnosis with the date of the first qualifying EHR diagnosis defined as the index date.

### Injury severity

Large-scale EHR databases do not commonly capture canonical measures of TBI (e.g., Glasgow Coma Scale). Therefore, injury severity was estimated using head or face Abbreviated Injury Score (AIS)^30^ values mapped to ICD codes using the ICD Programs for Injury Categorization (ICDPIC) package;^31^ values were then stratified by less serious (AIS<3) or serious (AIS≥3).^32^ This approach has been previously utilized by EHR-based trauma and TBI studies.^32–34^

### Clinical and demographic measures

Self-reported sex, race, and ethnicity information was derived from the AoU “The Basics Survey”.^35^ Sex was categorized into female, male, other (intersex and “none of these”), and unknown (“skip” or “prefer not to answer”) groups. Race and ethnicity categories were combined into previously described groups:^36,37^ non-Hispanic Black or African American (NHB), non-Hispanic White (NHW), Hispanic, and multiracial. Remaining race-ethnicity groups were combined into an “all other races” category to account for limitations with sample size; these include American Indian or Alaskan Native (AIAN), Asian, Middle Eastern or North African (MENA), and Native Hawaiian or Other Pacific Islander (NHOPI). Unknown race-ethnicity represented the following survey responses: “none of these”, “skip”, or “prefer not to answer”. Area-based socioeconomic status was estimated using the Social Deprivation Index (SDI),^38^ a composite measure derived from seven characteristics of the American Community Survey.^39^ The Cohen Perceived Stress Scale (PSS)^40^ was used to assess perceived stress and was available for a subset of participants. The PSS is a 14-item Likert-scale instrument with established stratification into low (0-13), moderate (14-26), or high (27-40) groups.

### Outcome measures

Neuropsychiatric diagnosis (NPD) clusters following index TBI were the primary outcome measures assessed. For each participant, ICD-9 and ICD-10-CM diagnosis codes were extracted from EHR data and mapped to phecodes^41^ using a modified implementation of the PheTK package.^42^ Ten broad phecode-derived categories [Table S2] were adapted from our previously established NPD clusters,^23^ with diagnosis requiring the presence of two or more qualifying ICD codes.^43^ Mortality was evaluated as a secondary outcome, using participant death information, curated from EHR and HealthPro reporting, available in AoU.

### Allostatic load biomarker selection

Relevant biomarkers were chosen based on established AL frameworks^24–26^ and data availability in AoU.^44^ Measures were curated using scripts adapted from the AoU *All by All: Curation of lab measurements phenotypes* Workspace.^45^ Non-interpretable or physiologically implausible values were removed and biomarker distributions were winsorized at 1% followed by 0-1 scale normalization. Model estimation included all participants enrolled in AoU with available EHR and laboratory data to maximize model stability. The per-participant mean was calculated for each marker using all measures recorded prior to index injury date (for the TBI cohort) or before a randomly selected EHR encounter (for remaining participants included in model estimation), thus generating a pre-index-specific score. Indicators with limited coverage and non-significant factor loadings were iteratively excluded based on modification indices until acceptable fit was achieved.^46,47^

### Confirmatory factor analysis

Our final model included body mass index (BMI), systolic and diastolic blood pressure (BP), blood HDL cholesterol, blood triglycerides, blood hemoglobin A1C (HbA1C), and C-reactive protein (CRP). The pALI construct was estimated with a single-factor confirmatory factor analysis (CFA) using the maximum likelihood estimator with robust standard errors in *lavaan* (v0.6.21).^48^ Acceptable model fit was determined by Comparative Fit Index (CFI) >.95; Root Mean Square Error Approximation (RMSEA) <.08; and Standardized Root Mean Square Residual (SRMR) <.08.^49^ Latent factor estimation including all available data with missing values handled through a full information maximum likelihood framework. Main analyses were restricted to participants with the three physical (BMI, systolic BP, and diastolic BP) and at least one blood measurement (“3+1” analysis). Sensitivity analyses were performed to evaluate model stability, measurement aggregation method, and participant exclusion [Supplemental Materials].

Measurement invariance (MI) was assessed with multi-group CFA across race-ethnicity and sex groups.^50^ Stepwise evaluation of multi-group models was performed by applying configural, metric, and scalar constraints to each model. Given the large sample size, invariance was evaluated by iterative changes in fit indices (*Δ*CFI < .01 and *Δ*RMSEA <.015) rather than *_χ_*^2^ statistic.^51,52^ If change in fit indices exceeded the pre-specified cut-offs, partial measurement invariance was determined by iteratively releasing individual constraints. Race-ethnicity MI was restricted to participants in NHB, NHW, Hispanic, multiracial, and other groups, while sex MI was limited to male and female participants to account for limitations in sample size.

### Analytical approach

Univariate differences in the continuous pALI construct were assessed using t-test (binary categorical), analysis of variance (categorical ≥2 levels), and Pearson’s correlation (continuous). The associations between pALI and TBI outcomes were evaluated using a time-to-event framework which excluded participants missing sex, race-ethnicity, or SDI data. Follow-up was initiated six-months after the index TBI across all outcomes assessed to account for acute post-injury related sequalae or previously undiagnosed pre-existing conditions.^2^

Mortality risk was assessed from six months following the index TBI to the time of participant censoring (last documented EHR encounter) or recorded death. Kaplan-Meier curves were generated to assess differences across pALI groups. Cox proportional hazards models were then fit, with the multivariable model adjusted for injury severity, age at index injury, sex, race-ethnicity, and SDI. The proportional hazards assumption was evaluated using Schoenfeld residuals, and the final model combined the “other” and Multiracial race/ethnicity categories due to low frequencies and unstable coefficient estimates.

The analytical sample for each NPD cluster excluded participants with a pre-existing diagnosis of the corresponding incident outcome. Incident NPDs were examined using cumulative incidence functions and competing-risk regression models (Fine and Gray method) with death as a competing event. Models were adjusted for injury severity, age at index injury, sex, race-ethnicity, SDI, and pre-existing comorbid NPDs. Presence of pre-existing comorbid NPDs was included with a single binary covariate representing the presence of any associated comorbidity identified in univariable analyses (significance threshold *P*<.10). When applicable, an FDR correction for multiple comparisons was applied (*P_FDR_*). Analyses were performed in the All of Us Researcher Workbench using R (version 4.5.0) and Python (version 3.10.16).

## RESULTS

### TBI cohort characteristics

There were 4,552 participants with TBI who had sufficient blood-based and physical measurement data to estimate pALI [Table 1]. The majority of TBI diagnoses were classified as less serious (AIS<3, 66.3%, *n*=3,019) and the average age at index injury was 55.4±15.6 standard deviation (SD) years. A slightly higher proportion of the cohort were female (52.5%, *n*=2,388) compared to male (46.5%; *n*=2,116). The majority of participants were of NHW identity (62.1%, *n*=2,829), followed by NHB (17.5%, *n*=796), Hispanic (9.6%, *n*=435), and multiracial (7.7%, *n*=350).

**Table 1.** Cohort characteristics.

| Characteristic | N | Count (%) |
| --- | --- | --- |
| ICDPIC AIS severity | 4552 |  |
| Less serious (<3) |  | 3019 (66.3) |
| Serious ($\geq 3$) | | 1533 (33.7) |
| Age at index TBI, y | 4552 |  |
| mean ± SD | | 55.4±15.6 |
| Sex | 4504 |  |
| Female |  | 2388 (52.5) |
| Male |  | 2116 (46.5) |
| unknown <sup>a</sup> |  | 48 (1.1) |
| Race-ethnicity | 4552 |  |
| NH-Black |  | 796 (17.5) |
| NH-White |  | 2829 (62.1) |
| Hispanic |  | 435 (9.6) |
| multiracial |  | 350 (7.7) |
| All other races <sup>b</sup> | 142 |  |
| AIAN |  | 59 (1.3) |
| Asian |  | 61 (1.3) |
| MENA |  | <20 <sup>c</sup> |
| NHOPI |  | <20 <sup>c</sup> |
| Social deprivation index | 4300 |  |
| mean ± SD | | 0.3±0.1 |
| unknown |  | 252 (5.5) |
| Cohen's PSS | 1518 |  |
| Likert-scale, mean ± SD | | 16.0±8.2 |
| Low |  | 616 (13.5) |
| Moderate |  | 732 (16.1) |
| High |  | 170 (3.7) |
| unknown |  | 3034 (66.7) |

<sup>a</sup> Includes participants who responded with "skip", or "prefer not to answer".
<sup>b</sup> Race-ethnicity groups included within the other classification for subsequent analyses.
<sup>c</sup> No group may be presented with participant or incident counts less than 20 (0 is acceptable), in accordance with the All of Us Data and Statistics Dissemination Policy.
**Abbreviations:** TBI = traumatic brain injury; ICDPIC AIS = International Classification of Diseases Program for Injury Categorization head or face Abbreviated Injury Score; SD = standard deviation; AIAN = American Indian or Alaskan Native; MENA = Middle Eastern or North African; NHOPI = Native Hawaiian or other Pacific Islander; and PSS = Perceived Stress Scale.

### Estimation of pre-injury allostatic load index

The pre-injury physiological burden of cumulative stress was estimated with a single-factor latent pALI variable across all participants with available pre-index data (*n*=126,431). The final model demonstrated acceptable fit indices (*CFI*=.988, *RMSEA*=.035, *SRMR*=.028) with residual covariance constraints reflecting the correlation between blood pressure measures and the shared variance among lipid and glycemic markers [Figure 2A]. Construct equivalence across demographic characteristics was assessed using multi-group CFA. Models fit across sex (*n*=125,095) and race-ethnicity (*n*=122,777) groups both achieved partial scalar-invariance, demonstrating acceptable fit after releasing select intercept constraints [Tables S3 and S4].

**Figure 2.**
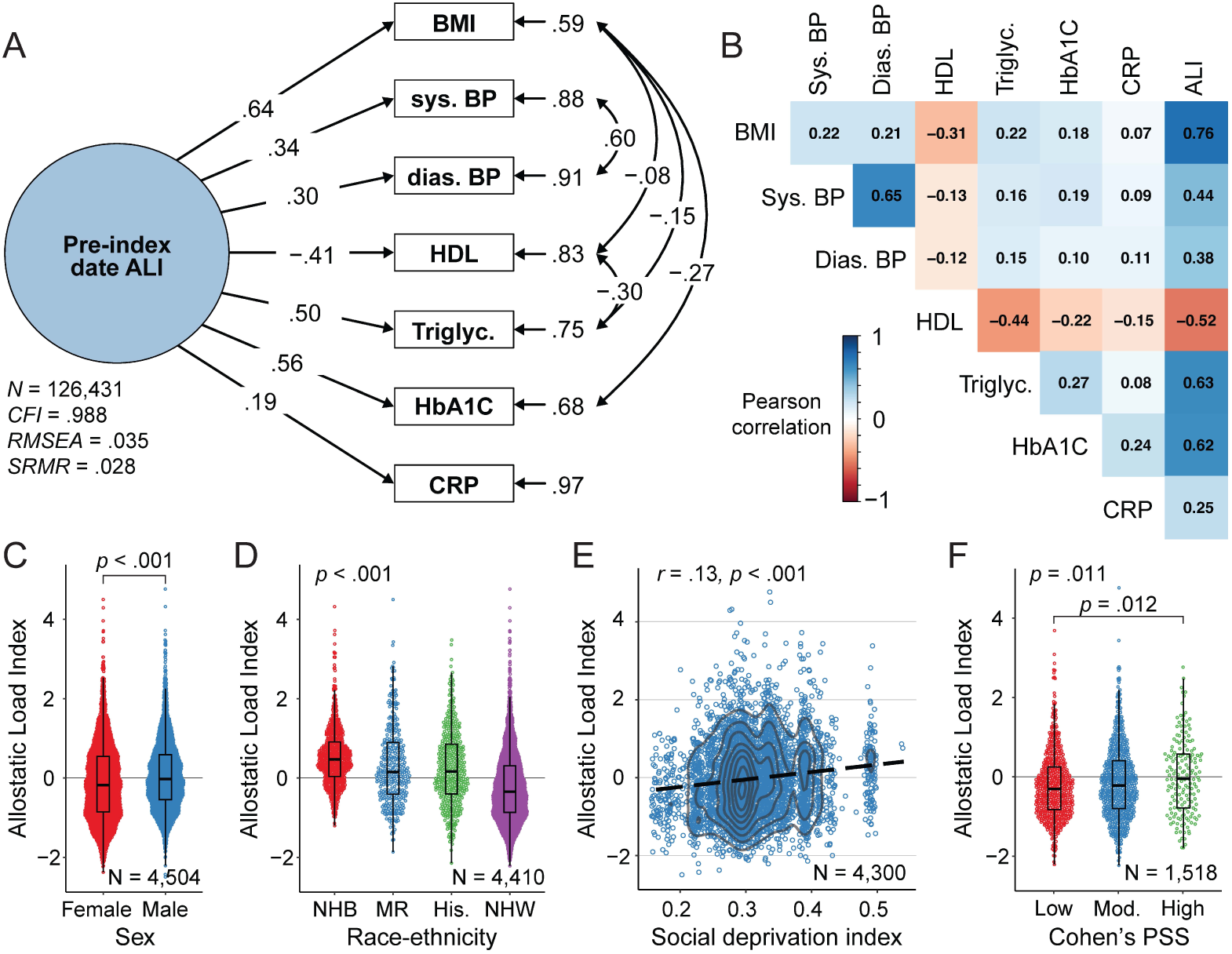
Pre-injury allostatic load index (pALI) estimation in All of Us. **(A)** pALI was estimated using single factor confirmatory factor analysis. Covariances across indicators were included based on model fit and physiological relevance. **(B)** Pearson’s correlation between the final pALI latent factor (race-ethnicity measurement invariant) and each pre-index input indicator. pALI scores were then compared across sociodemographic characteristics including **(C)** sex and **(D)** race-ethnicity groups. Race-ethnicity groups represented include non-Hispanic Black (NHB), NH-White (NHW), multiracial (MR), and Hispanic. **(E)** Pearson’s correlation between pALI and social deprivation index. **(F)** Comparison of ALI across Cohen’s Perceived Stress Score (PSS) cut-offs: low (0-13), moderate (mod.; 14-26), and high (27-40). Comparison across categorical variables performed with Welch’s two-sample t-test or ANOVA and Tukey-t post-hoc, where applicable.

Variation in AL has been previously observed across racial and ethnic groups;^11,53^ therefore, latent factors values estimated from the race-ethnicity multi-group CFA were used as the primary measure of pALI in main analyses. The final construct demonstrated moderate to strong correlation with all pre-index input indicators (*r-*absolute ≥ .25). Correlations between the latent factor and its constituent indicators exceeded the pairwise correlations observed among the indicators themselves, except for between systolic and diastolic blood pressure (*r*=.65) [Figure 2B]. For descriptive analyses, pALI was grouped into low (−3.1 ≥ pALI ≤ −.51), moderate (−.51 > pALI ≤ .36), and high (.36 > pALI ≤ 6.70) categories [Table S5].

### Differences in pre-injury allostatic load across sociodemographic characteristics

Variability in pALI was first assessed across sociodemographic characteristics. Male participants were observed to have a marginally higher pALI (mean=.08), compared to the female group (mean=−.10, *P*<.001) [Figure 2C]. A significant difference in pALI was also observed across race-ethnicity groups (*P*<.001), with the NHW-group observed to have the lowest pALI (mean=−.22), while the NHB-group had the highest (mean=.53) [Figure 2D]. pALI was positively correlated with area-level SDI (*r*=.13, *P*<.001) [Figure 2E]. Cohen’s PSS was available for a subset of individuals in the TBI cohort (33.3%, *n*=1,518) [Table 1] and was positively correlated with pALI (*r*=.08, *P*=.002). A significant difference in pALI was observed across established PSS score cut-points *(P=.*011), which was driven by the difference between the highest (ALI mean=0.01) and lowest (ALI mean=−.22, *P*=.012) PSS groups [Figure 2F].

### Association between pre-injury allostatic load and post-TBI mortality

Allostatic load has been previously associated with increased risk of neurological morbidity as well as mortality.^12,54,55^ To assess how this conferred risk may be represented following brain trauma, we assessed the association between pALI and post-TBI mortality risk. A significant difference in mortality rate was observed across survival curves stratified by low, moderate, and high pALI groups (Log-Rank test *P*=.008) [Figure 3A]. Cox proportional hazards models were then fit [Figure S1] and demonstrated a significant association between higher pALI and an increased hazard ratio (HR) of mortality in crude (HR=1.49 [95%CI, 1.22-1.82]; *P*<.001) and adjusted (adjusted HR [AHR]=1.71 [95%CI, 1.36-2.14]; *P*<.001) models [Table S6]. Models stratifying pALI by quantile demonstrated an adjusted post-TBI mortality hazard ratio 4.15-times higher (95%CI, 1.37-12.61; *P*=.012) in the highest (pALI >1.23 and ≤ 6.70) compared to the lowest (pALI ≥ −3.10 and ≤ −1.16) quantile [Figure 3B and Figure S2].

**Figure 3.**
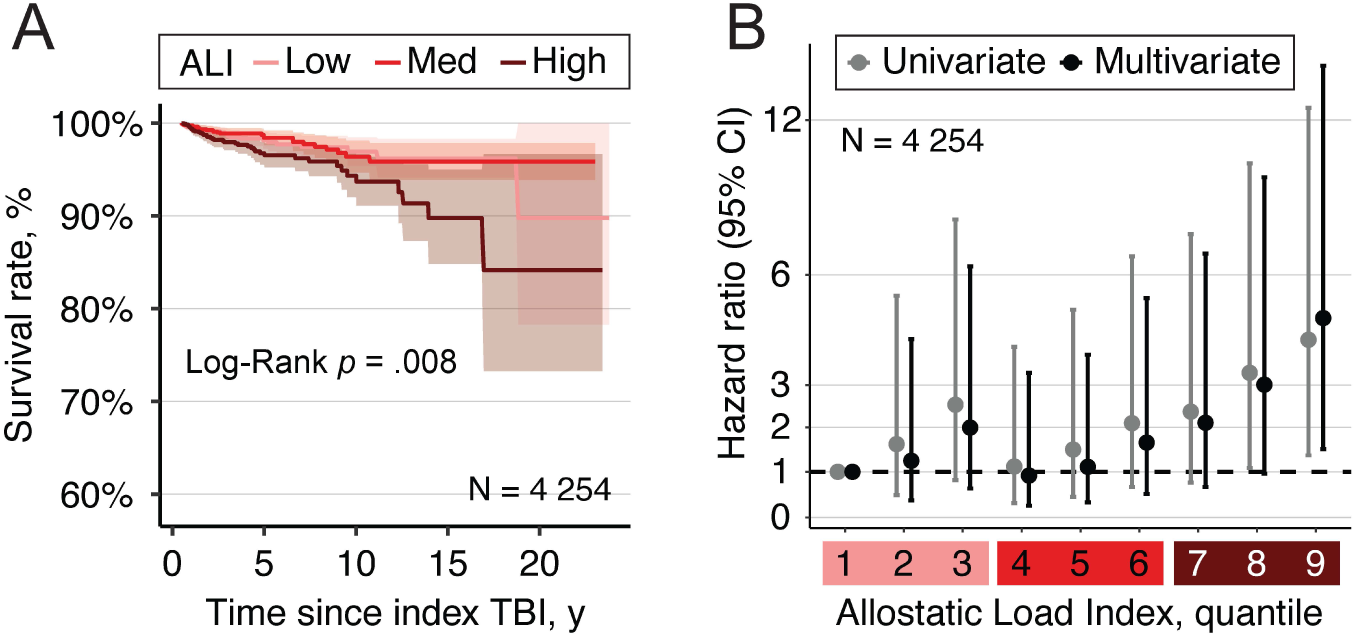
Association between pre-injury allostatic load index (pALI) and post-TBI mortality. **(A)** Kaplan-Meier curve assessing post-TBI mortality stratified by low, medium (med), and high pALI group. **(B)** Cox-proportional hazards models assessing association between post-TBI mortality risk across pALI quantiles. Colored boxes surrounding pALI quantiles correspond to pALI groups (low, medium, high).

### Pre-injury allostatic load is associated with post-TBI incident neuropsychiatric outcomes

The association between pALI and incident post-TBI NPD was then examined while accounting for the observed relationship between pALI and post-injury mortality. The cumulative incidence of each NPD [Table S7] was estimated and compared across low, moderate, high pALI groups with significant differences for post-TBI PTSD (Gray’s test *P*=.002) and sleep disorder (Gray’s test *P*<.001) diagnoses, while post-TBI mood disorder diagnoses trended towards significance (Gray’s test *P*=0.085) [Figure S3].

Competing-risk regression models were fit for each NPD cluster to assess the association with pALI after adjusting for clinical and demographic factors. Higher pALI was associated with increased HR of post-TBI PTSD and sleep disorders [Figure 4]. pALI was associated with increased risk of incident PTSD diagnosis in both crude (HR=1.33 [95%CI, 1.14-1.56]; *P_FDR_*=.001) and adjusted (AHR=1.28 [95%CI, 1.10-1.50]; *P_FDR_*=.009) models. A similar pattern was observed for sleep disorder diagnoses, with elevated risk (crude HR=1.32 [95%CI, 1.20-1.45]; *P_FDR_*<.001) maintained after model adjustment (AHR=1.42 [95%CI, 1.29-1.57]; *P_FDR_*<.001). pALI was associated with higher risk of mood disorder diagnosis in the univariate model (HR=1.21 [95%CI, 1.07-1.37]; *P_FDR_*=.011), though was no longer significant in the adjusted model after correction for multiple comparisons (AHR=1.17 [95%CI, 1.02-1.34]; *P_FDR_*=.10). The associations between pALI and post-TBI PTSD and sleep disorder diagnoses remained directionally consistent across sensitivity analyses [Supplemental Materials and Figure S4-S6]; however, these analyses also suggested that the most stable pALI construct relied on the full set of contributing indicators.

**Figure 4.**
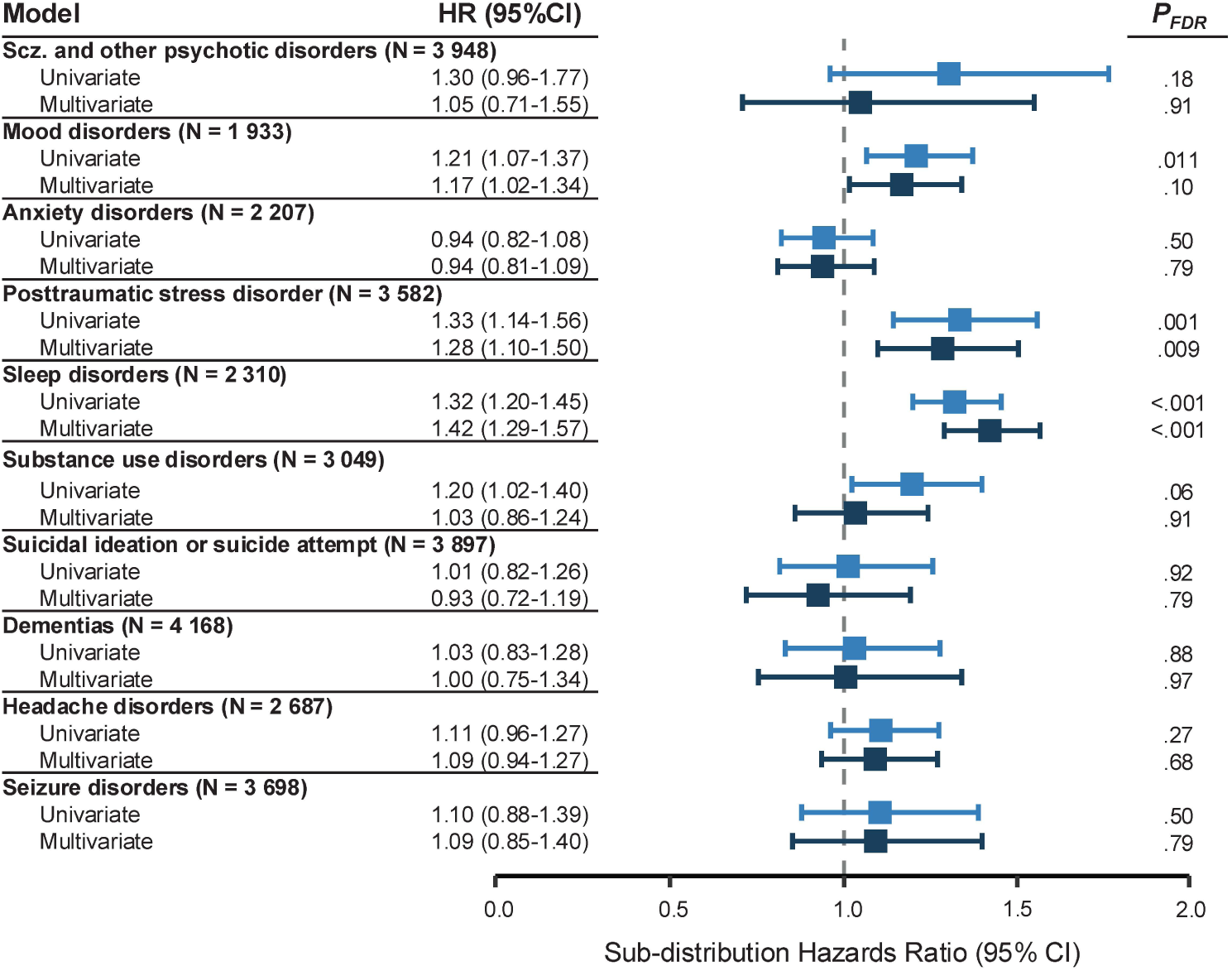
Competing risk model pre-injury ALI (pALI) regression coefficients. Univariable and multivariable pALI coefficients for each neuropsychiatric diagnostic (NPD) cluster displayed as the sub-distribution hazard ratio (HR) with 95% confidence intervals. Counts next to each NPD represent the total *N* at risk for each model. *P* values displayed have been corrected for multiple comparisons (FDR).

## DISCUSSION

Higher pre-injury AL was associated with increased risk incident neuropsychiatric diagnoses and mortality following TBI, supporting our hypothesis that pre-injury cumulative stress burden may modify longer term post-injury outcomes. These findings emphasize the need for multidimensional characterization of TBI, as outlined in the CBI-M framework,^6^ which positions TBI not as an isolated neurological event, but as an acute insult occurring against a background of physiological compromise. Within this framework, AL may serve as a unifying model that indexes the physiological consequences of chronic stress exposure, including both generalized risk and specific vulnerability for neuropsychiatric outcomes.

The selective associations observed between pALI and incident PTSD and sleep disorders may suggest an overlapping stress-sensitivity phenotype. PTSD and sleep-wake disorders are highly comorbid with TBI,^56,57^ and occur at elevated rates following injury.^58,59^ Co-occurrence of these disorders has been proposed to reflect shared hyperarousal, neuroendocrine dysregulation, and altered HPA-axis signaling^60,61^ – processes closely linked to stress physiology.^16^ Furthermore, pathways implicated in both PTSD and sleep disorders overlap with mechanisms involved in secondary injury cascades, including neuroimmune activation,^62,63^ glutamate excitotoxicity,^64^ and oxidative stress.^3^ Elevated pALI may capture stress-sensitive vulnerabilities that reinforce neuroinflammation and impaired recovery, contributing to disturbances in arousal regulation, sleep continuity, and trauma-related symptom development following TBI. This pattern is also consistent with the concept of “operator syndrome”, described in military and tactical populations exposed to repeated traumatic and high-stress environments,^65^ providing a parallel for how cumulative stress burden may manifest through overlapping neuropsychiatric phenotypes.

Our findings also highlight the role of AL in biologically contextualizing the CBI-M Modifier pillar^4^ by operationalizing the physiological burden of social disadvantage. We observed significant differences in pALI across sociodemographic characteristics that pattern exposure to psychosocial stress,^7,53^ including higher AL among NHB participants and those with greater social deprivation, suggesting unequal stress burden across groups. These associations parallel established disparities in TBI, with NHB individuals experiencing worse TBI prognosis and higher incident neuropsychiatric outcome risk, while socioeconomic disadvantage is associated with worse recovery.^22,66,67^ Structural inequities – including social deprivation, structural racism, and unequal access to social and healthcare services – may contribute, in part, to differences in TBI outcome by imposing chronic psychosocial stress that accumulates over the lifetime as AL; these effects may manifest through biological “weathering”, neurological morbidity, and elevated mortality risk.^8,11,53,68^ Together, pre-injury exposure to psychosocial stress emerges as one pathway through which BPSE factors may modify post-TBI trajectory.

### Limitations

Our analyses relied on retrospective EHR-linked data, which are subject to biases inherent to observational research, including diagnostic code misclassification, variable missingness, and biases related to healthcare engagement. TBI and subsequent neuropsychiatric condition diagnoses may be subject to selection processes that impact ascertainment and EHR capture.^22^ Heterogeneity in healthcare utilization and laboratory testing likely influenced the availability and timing of component indicators; for example, CRP is often obtained in specific clinical contexts, e.g., suspected infection or inflammatory illness, rather than during routine exam. Finally, while the pALI conceptually captures pre-injury stress exposure, component indicators may also individually represent underlying health conditions not directly related to stress.

## CONCLUSIONS

Our findings suggest that the biological consequences of chronic stress exposure may represent an underrecognized modifier of post-TBI trajectory. Under the central framework through which BPSE factors exert measurable psychosocial stress burden, AL may serve as a centrally linking mechanism connecting environmental stress, chronic disease, and heterogeneous outcomes in TBI. These findings support the incorporation of stress-related measures into multidimensional models of TBI classification and underscore the need for future studies to investigate how stress vulnerability and resilience shape neurological health.

## Data Availability

All data produced in the present study are available upon reasonable request to the authors. Original data are available to researchers who complete training through the All of Us Research Program (https://www.researchallofus.org/). The workspace used for the analysis presented in this study can be found in the Research Project Directory (https://www.researchallofus.org/research-project-directory/) and is named "Role of chronic stress on neuropsychiatric sequelae following TBI (v8)".

## ACKNOWLEDGMENTS

The authors would like to thank participants for their willingness to participate in the AoU Research Program and contribute their data. We also would like to acknowledge the National Institutes of Health All of Us Research Program for making the participant data available for this study.

## AUTHORSHIP CONTRIBUTION STATEMENT (CREDIT FORMAT)

Conceptualization: T.H.W., T.B.B., and E.J.B.

Data curation: T.H.W.

Formal analysis: T.H.W.

Funding acquisition: T.B.B. and E.J.B.

Investigation: T.H.W.

Methodology: T.H.W., P.B.B., T.B.B., and E.J.B.

Project administration: T.B.B., and E.J.B.

Resources: T.B.B. and E.J.B.

Software: T.H.W. and P.B.B.

Supervision: P.B.B., T.B.B., and E.J.B.

Validation: T.H.W.

Visualization: T.H.W.

Writing – original draft: T.H.W.

Writing - review & editing: T.H.W., P.B.B., T.B.B., and E.J.B.

## FUNDING STATEMENT, EVEN WHEN NOT APPLICABLE

This work was supported by the National Institutes of Health Clinical Research Scholars Training (CREST) Program Grant (5R25MD017950-02).

