## Supplemental Materials for "Allostatic load modifies neuropsychiatric risk following traumatic brain injury"

#### Sensitivity Analysis Methods

Sensitivity analyses were performed to evaluate (1) CFA model stability, (2) the impact of alternative pre-index measure aggregation approaches, and (3) the potential effect of participant exclusion due to data missingness. To assess latent factor stability and the relative contribution of individual indicators, the pre-injury allostatic load index (pALI) factor was re-estimated under a leave-one-out (LOO) framework; one marker at a time was excluded from pALI estimation producing a new latent construct estimated from the remaining six indicators. Second, to determine how using the pre-injury average may impact the results, pALI was re-estimated using pre-index values aggregated by the median, minimum, and nearest to the index date value. Finally, the potential impacts of participant exclusion were assessed by estimating pALI with expanded inclusion criteria, which considered all participants with at least one ALI-related measure, compared to participants meeting the primary "3+1" criterion which were restricted to participants who had the three physical measurements (BMI, systolic BP, and diastolic BP) as well as at least one blood measurement.

#### Sensitivity Analysis Results

We re-estimated the pALI latent construct under a leave-one-out (LOO) framework to evaluate model stability and determine if the observed associations were unduly driven by any single indicator. Across the seven LOO models, the direction of associations between pALI and NPD clusters remained consistent in both univariate and multivariable models [Figure S4]. Observed associations with PTSD and sleep disorders were preserved across all LOO models at nominal significance ($P$<.05); although, significance was not maintained after FDR correction in post-TBI PTSD models that excluded triglycerides (crude $P_{FDR}$=.09; adjusted $P_{FDR}$=.1) or HbA1C (crude $P_{FDR}$=.17; adjusted $P_{FDR}$=.11), and in the univariate model excluding diastolic BP (crude $P_{FDR}$=.06; adjusted $P_{FDR}$=.035). The nominal association observed in the adjusted mood disorder model was not maintained in multivariable LOO models the excluded BMI ($P$=.27), triglycerides ($P$=.07), and HbA1C ($P$=.09). These findings suggest that the observed main effects are stable; although, the strongest and most cohesive pALI construct relies on the full set of contributing indicators.

Next, the pALI construct was re-estimating using alternative per-participant measure aggregation approaches. Pre-index participant measures were re-aggregated by the median, minimum, or value closest to the index injury to determine how estimation using the per-participant mean may have impacted our results. The direction of associations was preserved across the different aggregation strategies, with nominal significance ($P$<.05) maintained across all models for PTSD and sleep disorder outcomes [Figure S5]. For PTSD, the multiple-comparisons threshold was not preserved the multivariate model aggregating measures by the minimum pre-index value (adjusted $P_{FDR}$=.11). The association between pALI and sleep disorders remained statistically significant across all aggregation approaches. The nominal adjusted effect of pALI on mood disorders was preserved for pALI estimated using the median (adjusted $P$=.033) or closest-to-index (adjusted $P$=.033) values, though was no longer significant in the model estimated using the minimum pre-index value (adjusted $P$=.10).

Finally, the potential impacts of data availability were assessed by re-estimating pALI with a more relaxed participant inclusion criteria, which included all participants with at least one pre-index measurement used to estimate pALI (total estimation sample *N*=223,549 with 6,114 in TBI cohort). The results for the associations between pALI and post-TBI PTSD or sleep disorders remained unchanged in both direction and statistical significance ($P_{FDR}$<.05) [Figure S6]. Interestingly, the association between pALI estimated from this more inclusive cohort and mood disorders met the multiple comparisons threshold in multivariable analysis ($P_{FDR}$=.003), suggesting that increased statistical power may be required to fully appreciate this observed effect.

### Supplemental Tables

**Table S1**

**ICD9/10-CM code definition for traumatic brain injury in All of Us.** Table is originally derived from Wroblewski *et al.* 2026 (doi: 10.1093/aje/kwaf030).

| Code | Description |
|---|---|
| **ICD9CM** | |
| 800 | fracture of vault of skull |
| 801 | fracture of the base of skull |
| 802 | Fracture of face bones |
| 803 | other and unqualified skull fractures |
| 804 | multiple fractures involving skull or face with other bones |
| 850 | concussion |
| 851 | cerebral laceration and contusion |
| 852 | subarachnoid, subdural, and extradural hemorrhage, following injury |
| 853 | other and unspecified intracranial hemorrhage following injury |
| 854 | intracranial injury of other and unspecified nature |
| 854.1 | Intracranial injury of other / unspecified nature with open intracranial wound |
| 950.1 | injury to the optic nerve and pathways |
| 950.2 | Injury to optic pathways |
| 950.3 | Injury to visual cortex |
| **ICD10CM** | |
| *S02.X* | |
| S02.0 | Fracture of Vault of Skull |
| S02.1 | Fracture of Base of Skull |
| S02.3 | Fracture of orbital floor |
| S02.7 | Multiple fractures involving skull and facial bones |
| S02.8 | Fracture of Other Specified Skull and Facial Bones |
| S02.9 | Fracture of Unspecified Skull and Facial Bones |
| S02.91 | unspecified fracture of skull |
| S04.0 | injury to optic nerve and pathways |
| S06 | Intracranial injury (all of S06) |
| *S07.X* | |
| S07.0 | Crushing injury of face |
| S07.1 | Crushing injury of skull |
| S07.8 | Crushing injury to other parts of head |
| S07.9 | Crushing injury of head, part unspecified |
| T02.0 | Fractures involving head with neck |
| T04.0 | Crushing injuries involving head with neck |
| T06.0 | Other injuries involving brain, cranial nerves, and spinal cord at neck level |

**Table S2**

**Neuropsychiatric diagnostic clusters derived from phecodes.** Outcomes were groups into 10 diagnostic clusters based on broad phecode categories. Condition descriptions represent the definition for each phecode included. Table adapted from Wroblewski *et al.* 2026 (doi: 10.1093/aje/kwaf030).

| Group | Phecodes | Descriptions |
|---|---|---|
| Schizophrenia and other psychotic disorders | 295; 295.1; 295.2; 295.3 | Schizophrenia and other psychotic disorders; Schizophrenia; Paranoid disorders; Psychosis |
| Mood disorders | 296; 296.1; 296.2; 296.22 | Mood disorders; Bipolar; Depression; Major depressive disorder |
| Anxiety disorders | 300; 300.1; 300.11; 300.12; 300.13; 300.2; 300.3; 300.4; 300.8 | Anxiety disorders; Anxiety disorder; Generalized anxiety disorder; Agoraphobia, social phobia, and panic disorder; Phobia; Generalized anxiety & phobic disorders; Obsessive-compulsive disorders; Dysthymic disorder; Acute reaction to stress |
| Post-traumatic stress disorder | 300.9 | Post-traumatic stress disorder |
| Sleep disorders | 327; 327.1; 327.3; 327.31; 327.32; 327.4; 327.41; 327.5; 327.6; 327.7; 327.71; 327.72 | Sleep disorders; Hypersomnia; Sleep apnea; Central/nonobstructive sleep apnea; Obstructive sleep apnea; Insomnia; Organic or persistent insomnia; Parasomnia; Circadian rhythm sleep disorder; Sleep related movement disorders; Restless legs syndrome; Sleep related leg cramps |
| Substance use disorders | 290.2; 316; 316.1; 317; 317.1; 317.11 | Delirium due to conditions classified elsewhere; Substance addiction and disorders; Polyneuropathy due to drugs; Alcohol-related disorders; Alcoholism; Alcoholic liver damage |
| Suicidal ideation or suicide attempt | 297; 297.1; 297.2; 969 | Suicidal ideation or attempt; Suicidal ideation; Suicide or self-inflicted injury; Poisoning by psychotropic agents |
| Dementias | 290.1; 290.11; 290.12; 290.13; 290.16 | Dementias; Alzheimer's disease; Dementia with cerebral degenerations; Senile dementia; Vascular dementia |
| Headache disorders | 306.9; 339; 340; 340.1 | Tension headache; Other headache syndromes; Migraine; Migraine with aura |
| Seizure disorders | 345; 345.1; 345.11; 345.12; 345.3 | Epilepsy, recurrent seizures, convulsions; Epilepsy; Generalized convulsive epilepsy; Partial epilepsy; Convulsions |

**Table S3**

**Multi-group confirmatory factor analysis across sex groups assessing measurement invariance.** Performed across male and female participant groups (n=125,095).

| Test | Original | Configural | Metric | Scalar | Partial scalar [a] |
|---:|---|---|---|---|---|
| Chi-square | 1376.8 | 1780.6 | 2068.0 | 18173.0 | 2113.8 |
| *P*-Value (Chi-square) | $<2\times10^{-16}$ | $<2\times10^{-16}$ | $<2\times10^{-16}$ | $<2\times10^{-16}$ | $<2\times10^{-16}$ |
| CFI | 0.988 | 0.985 | 0.983 | 0.847 | 0.982 |
| TLI | 0.973 | 0.965 | 0.970 | 0.785 | 0.972 |
| RMSEA | 0.035 | 0.040 | 0.037 | 0.098 | 0.036 |
| SRMR | 0.028 | 0.029 | 0.032 | 0.086 | 0.031 |
| ΔCFI | N/A | -0.003 | -0.002 | -0.136 | <.001 |
| ΔRMSEA | N/A | 0.005 | -0.003 | 0.061 | -0.001 |

[a] The following intercept constraints were released across sex groups: BMI, triglycerides, HDL, and HbA1C.

**Table S4**

**Multi-group confirmatory factor analysis across race-ethnicity groups assessing measurement invariance.** Performed across NH-White, NH-Black, Hispanic, multiracial, and other race-ethnicity groups (n=122,777); participants with unknown race-ethnicity status were excluded from multi-group models.

| Test | Original | Configural | Metric | Scalar | Partial scalar [a] |
|---|---|---|---|---|---|
| Chi-square | 1376.8 | 1566.2 | 2390.9 | 10272.6 | 3183.3 |
| *P*-Value (Chi-square) | 0 | 0 | 0 | 0 | 0 |
| CFI | 0.988 | 0.987 | 0.980 | 0.912 | 0.973 |
| TLI | 0.973 | 0.969 | 0.969 | 0.900 | 0.963 |
| RMSEA | 0.035 | 0.037 | 0.037 | 0.067 | 0.041 |
| SRMR | 0.028 | 0.028 | 0.035 | 0.058 | 0.037 |
| ΔCFI | N/A | -0.001 | -0.007 | -0.068 | -0.007 |
| ΔRMSEA | N/A | 0.002 | <.001 | 0.030 | 0.004 |

[a] The following intercept constraints were released across race groups: triglycerides, HDL, diastolic and systolic blood pressure.

**Table S5**

**Grouping of pre-injury allostatic load index by quantile.** Allostatic load index quantiles were derived from the pre-injury race-ethnicity measurement invariant CFA; quantiles were further stratified into low, medium, and high pALI groups for descriptive analyses.

| ALI Group | Quantile | Pre-injury ALI | Count |
|---|---|---|---|
| Low | 1 | [-3.10,-1.16] | 438 |
| Low | 2 | (-1.16,-0.81] | 545 |
| Low | 3 | (-0.81,-0.51] | 494 |
| Medium | 4 | (-0.51,-0.23] | 539 |
| Medium | 5 | (-0.23,0.05] | 539 |
| Medium | 6 | (0.05,0.36] | 528 |
| High | 7 | (0.36,0.72] | 538 |
| High | 8 | (0.72,1.23] | 495 |
| High | 9 | (1.23,6.70] | 436 |

**Table S6**

**Cox proportional hazards models assessing association between pre-injury ALI and post-TBI mortality.** ALI was modelled as a continuous variable and effect sizes displayed at hazard ratios (HR) with 95% confidence intervals.

| **Variable** | **HR (95%)** | ***P*-Value** |
|---|---|---|
| pALI - Univariate | 1.49 (1.22-1.82) | <.001 |
| pALI - Multivariate | 1.71 (1.36-2.14) | <.001 |
| ICDPIC AIS severity (*ref*=less serious, <3) | | |
| Serious (≥3) | 2.51 (1.63-3.87) | <.001 |
| Age at index TBI, y | 1.06 (1.05-1.09) | <.001 |
| Race-ethnicity (*ref*=NH-Black) | | |
| NH-White | 1.19 (0.65-2.16) | 0.58 |
| Hispanic | 1.15 (0.52-2.55) | 0.72 |
| Other | 0.32 (0.04-2.41) | 0.27 |
| Sex (*ref*=female) | | |
| Male | 1.09 (0.71-1.69) | 0.69 |
| Social Deprivation Index | 2.67 (0.74-9.68) | 0.14 |

**Table S7**

**Counts for each neuropsychiatric diagnosis cluster assessed in competing risk analyses.** The total at risk represents all participants included in the model after excluding those with each respective NPD.

| NPD Cluster | N risk | N censored | N outcome | N deceased |
|---|---|---|---|---|
| Schizophrenia and other psychoses | 3948 | 3816 | 50 | 82 |
| Mood disorders | 1933 | 1656 | 248 | 29 |
| Anxiety disorders | 2207 | 1881 | 281 | 45 |
| PTSD | 3582 | 3348 | 158 | 76 |
| Sleep disorders | 2310 | 1826 | 443 | 41 |
| SUD | 3049 | 2849 | 150 | 50 |
| Suicidal ideation or attempt | 3897 | 3724 | 91 | 82 |
| Dementias | 4168 | 4024 | 62 | 82 |
| Headache disorders | 2687 | 2373 | 253 | 61 |
| Seizure disorders | 3698 | 3522 | 106 | 70 |

### Supplemental Figures

#### Figure S1

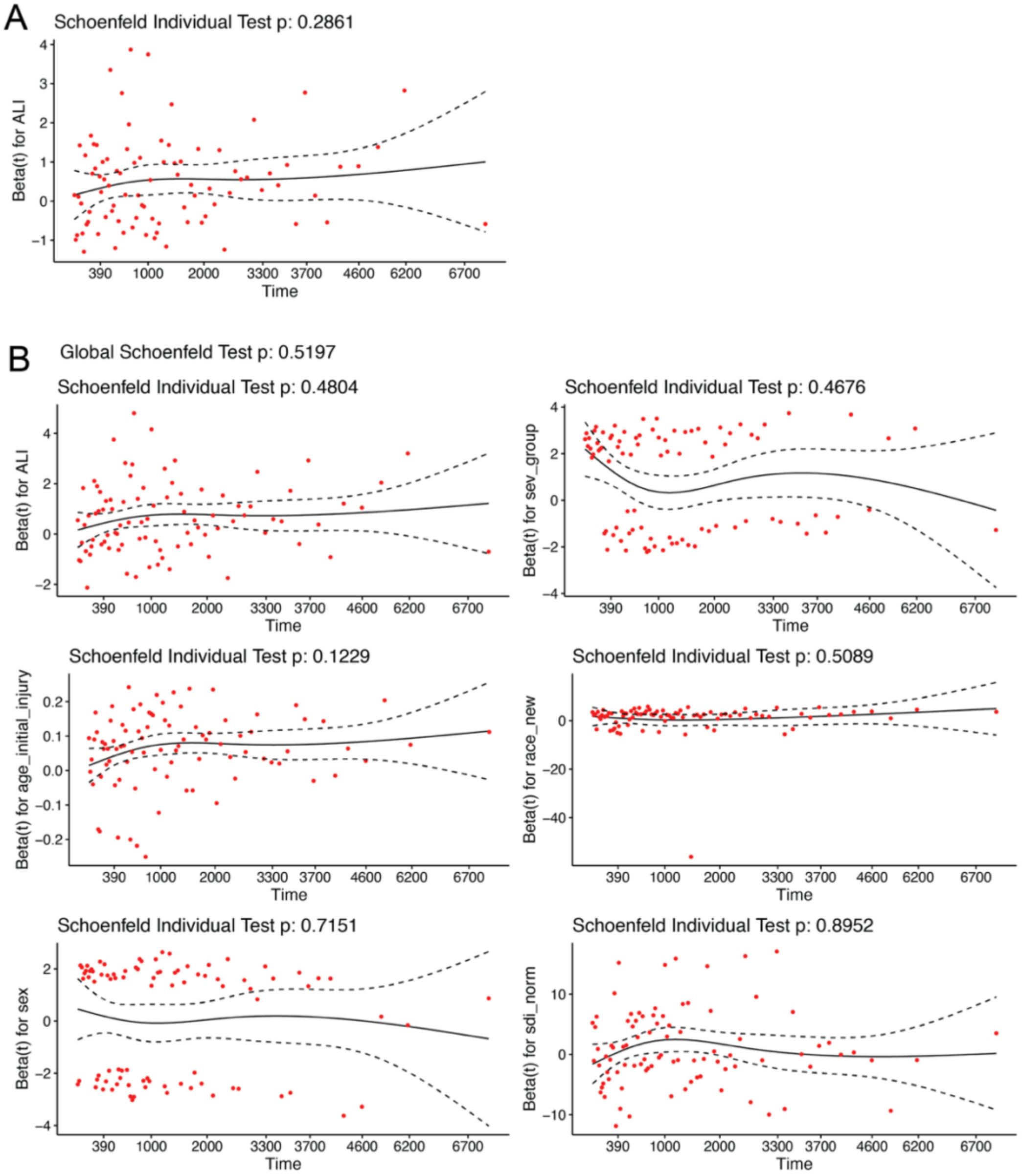

**Schoenfeld residuals for cox proportional hazards models assessing association between pre-injury ALI (continuous) and post-TBI mortality.** Residuals displayed for **(A)** univariate and **(B)** adjusted models.

**Figure S2**

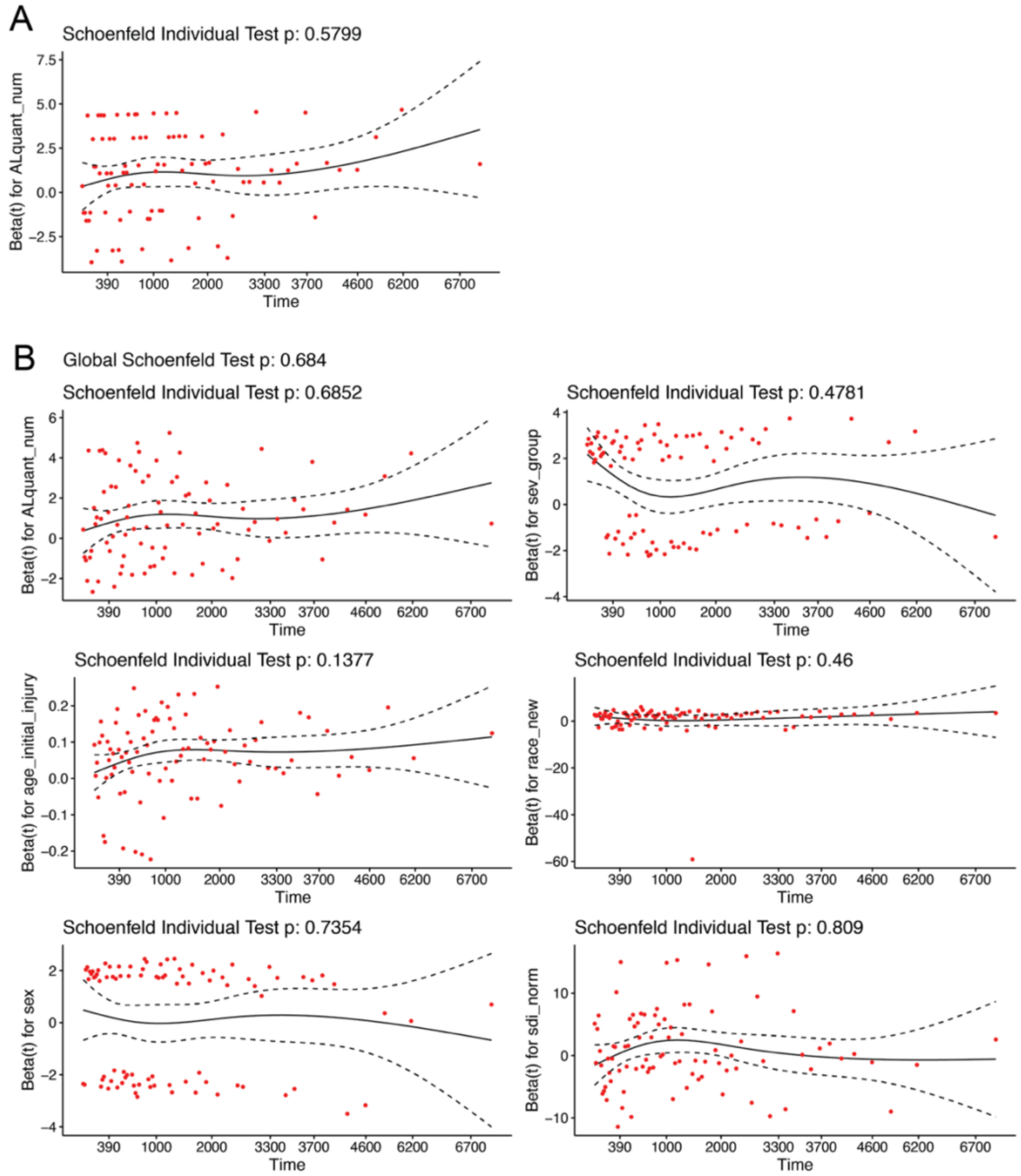

**Schoenfeld residuals for cox proportional hazards models assessing association between pre-injury ALI quantiles and post-TBI mortality.** Residuals displayed for **(A)** univariate and **(B)** adjusted models.

**Figure S3**

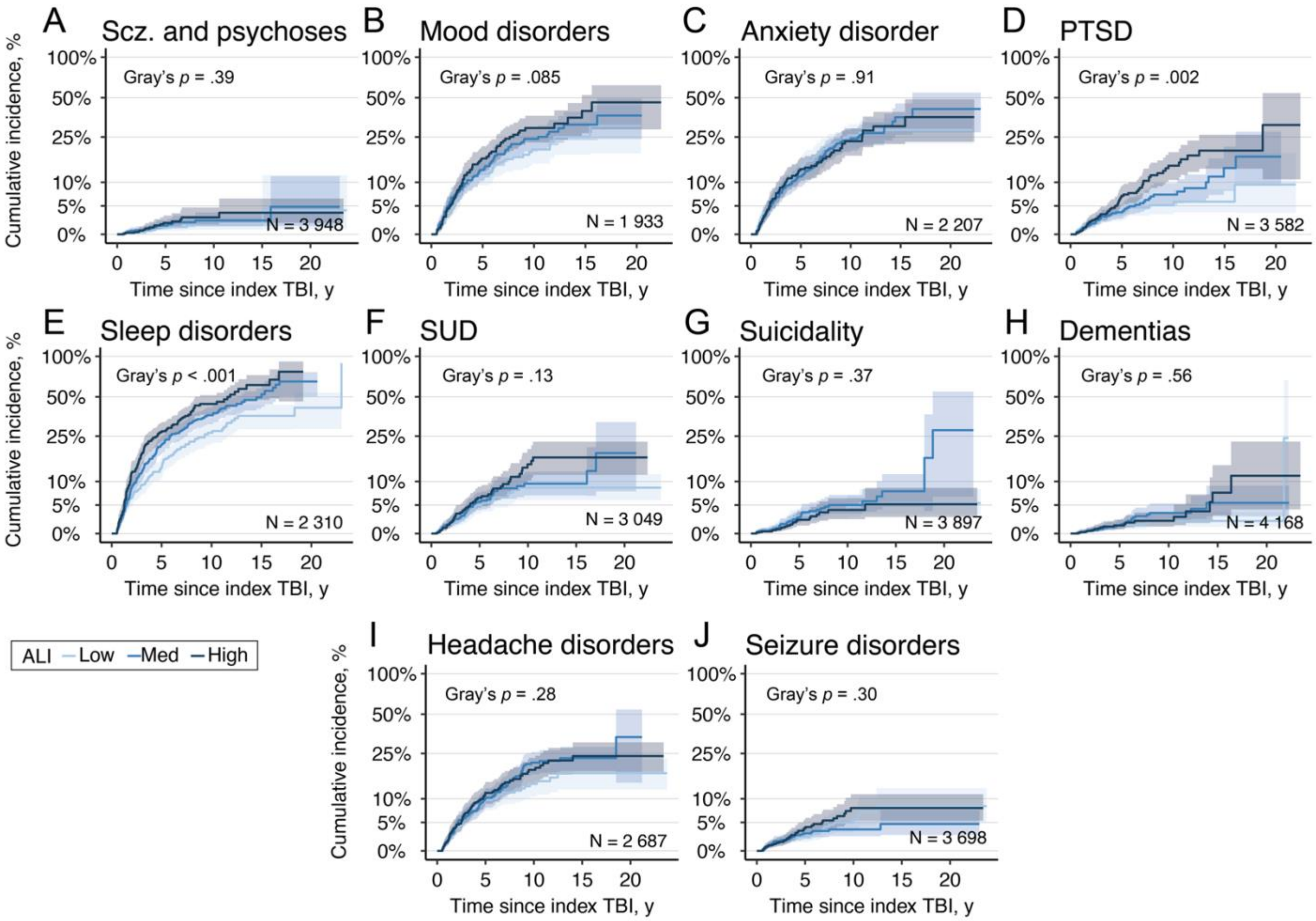

**Post-TBI neuropsychiatric diagnosis cumulative incidence functions stratified by pre-injury ALI (pALI) group.** Cumulative incidence was estimated with mortality as a competing risk and reported as percentage with 95% confidence interval ribbons. Differences in pALI groups was assessed using Gray's test.

**Figure S4**

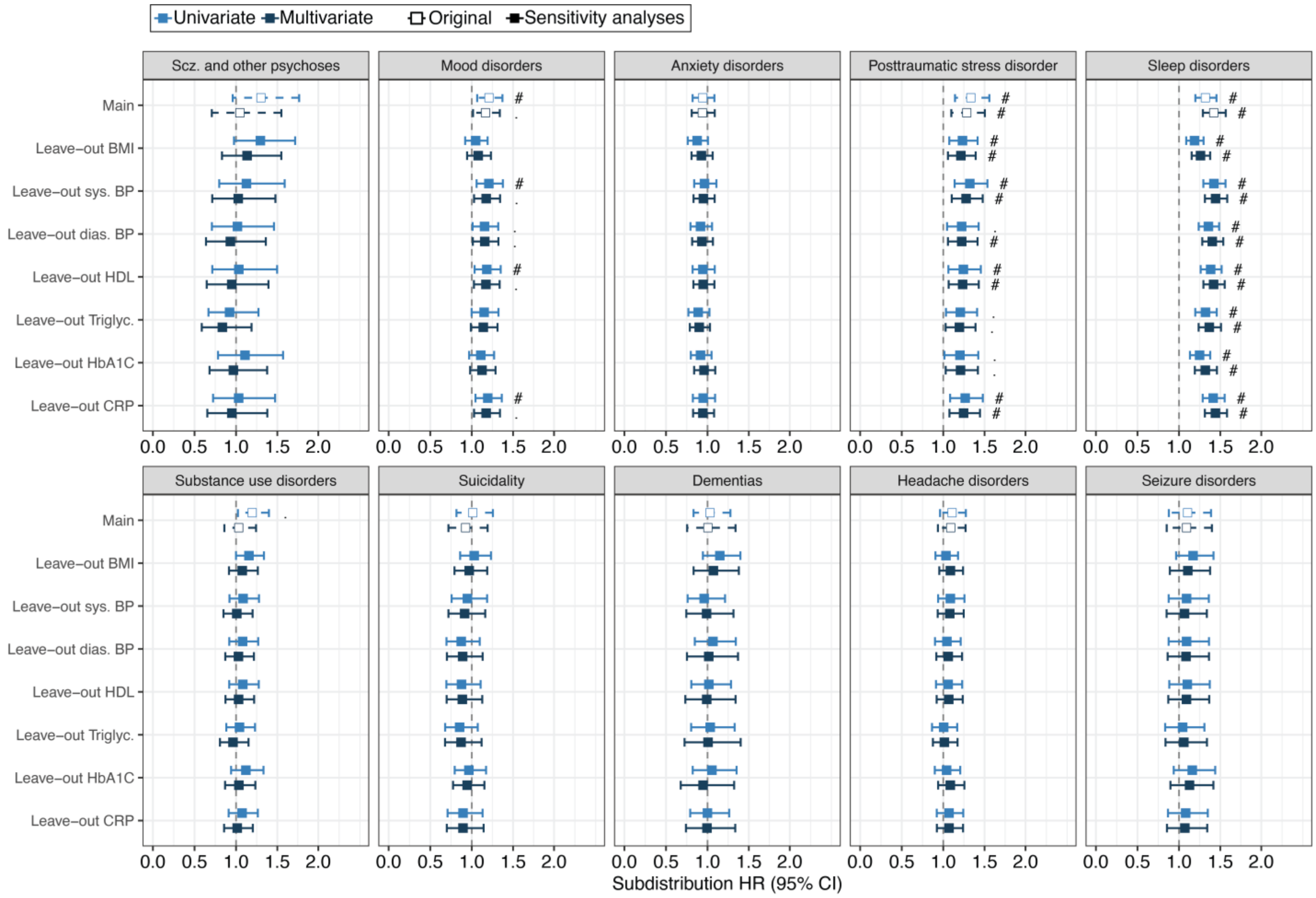

**Leave-one-out (LOO) pre-injury ALI (pALI) regression coefficients sensitivity analyses.** Univariable and multivariable competing risks regression coefficients for the main pALI construct and the seven LOO models faceted by neuropsychiatric diagnostic (NPD) cluster. Effects are displayed as the sub-distribution hazard ratio (HR) with 95% confidence intervals. Significance is displayed as nominal (*P*<.05; ".") and after correction for multiple comparisons ($P_{FDR}$<.05, "#").

**Figure S5**

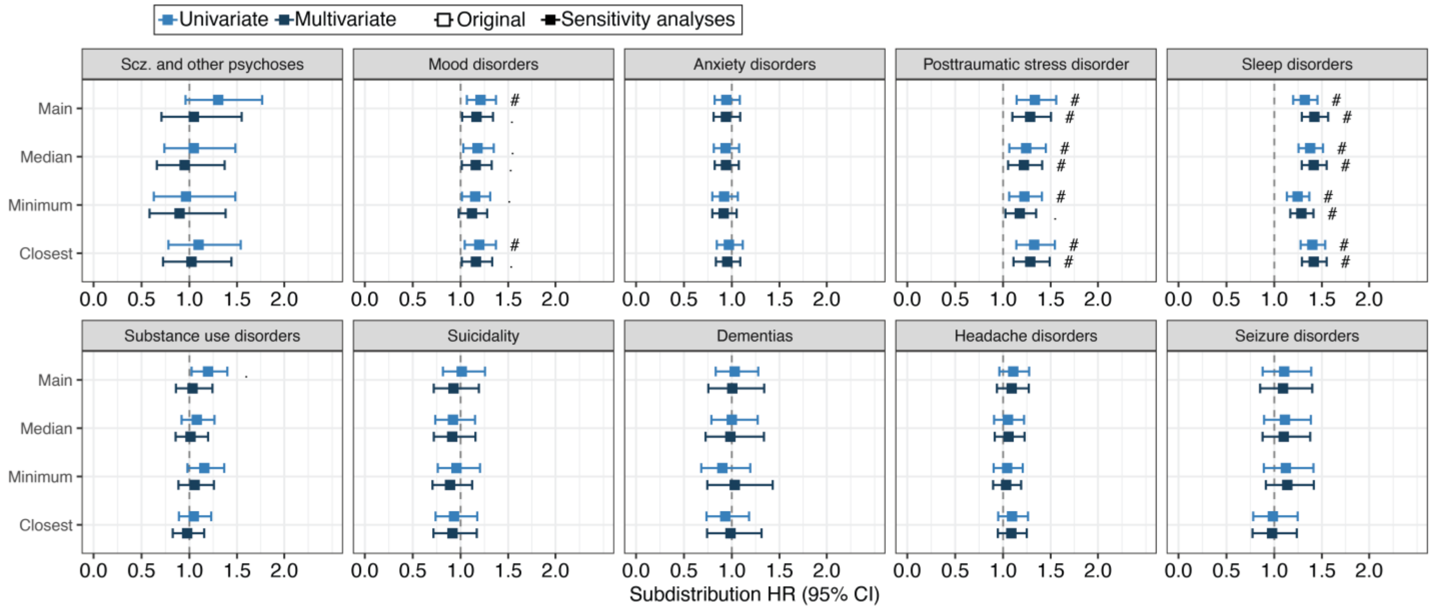

**Alternative pre-index measure aggregation sensitivity analyses.** Univariable and multivariable competing risks regression coefficients for the main pALI construct and pALI estimated from pre-injury median, minimum, and closest values for each indicator. Coefficients are stratified by neuropsychiatric diagnostic (NPD) cluster and displayed as the sub-distribution hazard ratio (HR) with 95% confidence intervals. Significance is displayed as nominal ($P$<.05; ".") and corrected for multiple comparisons ($P_{FDR}$<.05, "#").

**Figure S6**

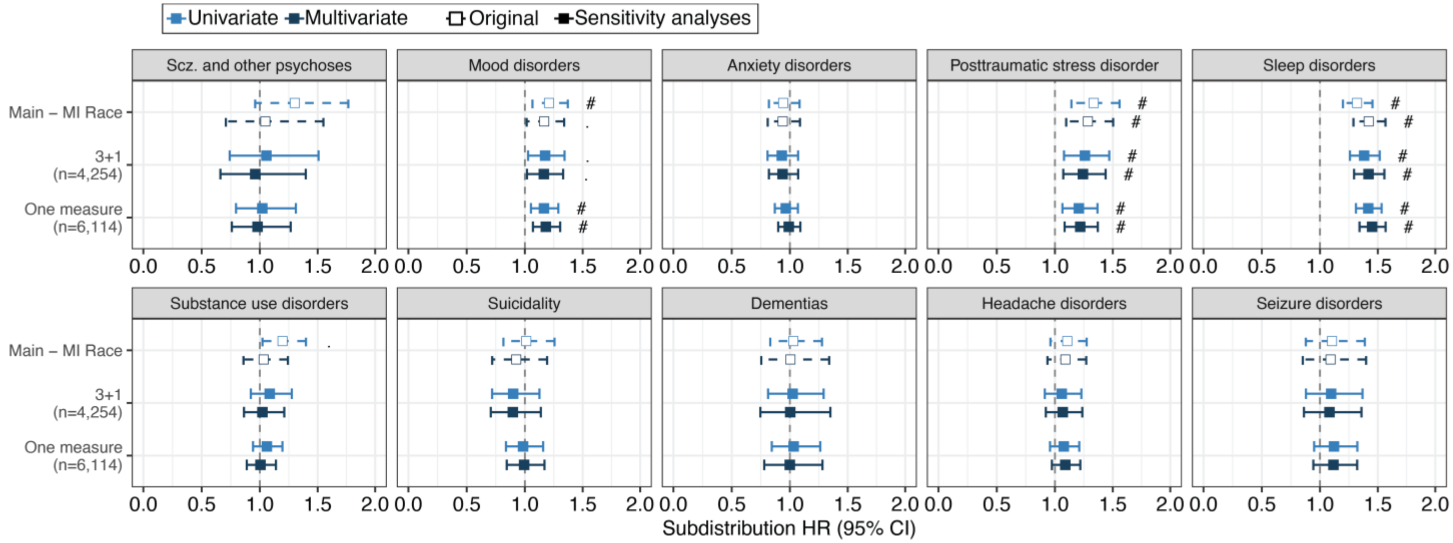

**Alternative participant inclusion criteria sensitivity analyses.** Univariable and multivariable competing risks regression coefficients for the main pALI construct and as well as the non-measurement invariant model (3+1) and model that only required one biomarker for participant inclusion. Coefficients are faceted by neuropsychiatric diagnostic (NPD) cluster with effects displayed as the sub-distribution hazard ratio (HR) with 95% confidence intervals. Significance is displayed as nominal (*P*<.05; ".") and after correction for multiple comparisons ($P_{FDR}$<.05, "#").
